# Leveraging Longitudinal Patient-Reported Outcomes Trajectories to Predict Survival in Non-Small-Cell Lung Cancer

**DOI:** 10.1101/2025.01.27.25321050

**Authors:** Jiawei Zhou, Benyam Muluneh, Zhaoyang Wang, Huaxiu Yao, Jim H. Hughes

## Abstract

**Purpose:** Despite their potential, patient-reported outcomes (PROs) are often underutilized in clinical decision-making, especially when improvements in PROs do not align with clinical outcomes. This misalignment may result from insufficient analytical methods that overlook the temporal dynamics and substantial variability of PROs data. To address these gaps, we developed a novel approach to investigate the prognostic value of longitudinal PRO dynamics in non-small-cell lung cancer (NSCLC) using Lung Cancer Symptom Scale (LCSS) data.

**Methods:** Longitudinal patient-reported LCSS data from 481 NSCLC participants in the placebo arm of a Phase III trial were analyzed. A population modeling approach was applied to describe PRO progression trajectories while accounting for substantial variability in the data. Associations between PRO model parameters and survival outcomes were assessed using Cox proportional hazards models. Model-informed PRO parameters were further used to predict survival via machine learning.

**Results:** A PRO progression model described LCSS dynamics and predicted a median time to symptom progression of 229 days (95% CI: 15-583). Faster PRO progression rates were significantly associated with poorer survival (HR 1.13, 95% CI: 1.076-1.18), while greater placebo/prior treatment effects correlated with improved survival (HR 0.93, 95% CI: 0.883-0.99). A machine learning model using PRO parameters achieved an AUC-ROC of 0.78, demonstrating their potential to predict overall survival.

**Conclusions:** This study demonstrates that longitudinal PRO data can provide prognostic insights into survival in NSCLC. The findings support the use of PRO dynamics to improve clinical decision-making and optimize patient-centered treatment strategies.

**Translational Relevance:** Patient-reported outcomes (PROs) offer a patient-centered, non-invasive approach to assessing health status and symptoms, making them a valuable tool for understanding the patient experience. However, PROs are often underutilized in clinical decision-making, especially when improvements in PROs do not align with clinical outcomes. This misalignment is often driven by analytical methods that fail to account for the temporal dynamics and inherent variability of PROs data. To address these limitations, we have developed a novel computational framework that integrates population modeling with machine learning to leverage longitudinal PRO dynamics for predicting survival in non-small-cell lung cancer patients. This innovative approach can be generalized to other PRO datasets, providing personalized insights into PRO trajectories for each patient. By enhancing the utility of PROs data, our framework holds the potential to significantly improve clinical decision-making, refine patient care strategies, and optimize the design and evaluation of clinical trials, ultimately advancing precision oncology.

## Introduction

Patient-reported outcomes (PROs) provide a patient-centered method for assessing health status and symptoms, offering systematic, non-invasive evaluations of physical and psychological well-being. (1,2) As direct assessments of patients’ experiences, PROs have been found to be independent predictors for survival outcomes in various types of cancer. (3,4)

Despite their potential, PROs are often underutilized in regulatory decision-making, particularly when improvements in PROs do not correspond to clinical outcomes.(5–7) In a survey based on oncology drugs approved between 2011 and 2017, only a quarter of them showed a statistically improvement in PROs.(8) Given the correlations between PROs and clinical outcomes, we hypothesize that the lack of statistical significance results from the limitations of traditional statistical methods in accurately analyzing PRO data. While previous studies have explored the associations between PROs and survival outcomes, most focused on baseline PRO values or changes at fixed time points, overlooking PRO dynamics during the study.(3,4,9) Notably, a recent study by Bhatt *et al*. in non-small-cell lung cancer (NSCLC) demonstrated that PRO dynamics are better correlated with tumor burden than single time-point measurements.(10) Furthermore, the PROs data have substantial variability due to its subjective nature. This large between-subject variability and intra-subject fluctuations can introduce noise into the data, reducing the statistical power needed to detect meaningful associations in traditional hypothesis tests.(11) Therefore, we aim to develop a novel computational method to address these challenges and leverage longitudinal PROs dynamics to predict patient survival.

Lung cancer remains the leading cause of cancer-related deaths globally, with over 125,070 deaths projected in 2024. (12) NSCLC accounts for 80-85% of lung cancer cases, with more than 30% patients diagnosed at stage III or IV. (13) NSCLC at advanced stages may cause a range of symptoms such as cough, pain, dyspnea, and fatigue, all of which dimmish patients’ QoL.(14) The Lung Cancer Symptom Scale (LCSS) is a well-established and reliable PRO tool for NSCLC. (15,16) This study analyzes longitudinal patient-reported LCSS data from the placebo arm of a Phase III NSCLC trial and assess the prognostic value of PRO dynamics for survival outcomes.

Given the inherent variability in PRO data, a population PRO progression model was developed to analyze the longitudinal PRO data for the entire study cohort simultaneously. This modeling approach allowed us to disentangle the large variability in PRO data into components such as inter-individual variability, measurement noise, and subpopulation heterogeneity. (17,18) The model successfully captured PRO trajectories for each NSCLC participant and was subsequently used to predict patient survival. The associations between PRO trajectories and clinical outcomes were further investigated, including overall survival (OS) and progression-free survival (PFS), and used model-informed PRO trajectories to predict patient survival through a machine learning approach. This framework demonstrates how longitudinal PRO data can enhance clinical decision-making and support the design of patient-centered treatment strategies in NSCLC.

## Materials and Methods

### Ethic statement

For this retrospective analysis, all data were de-identified and could not be traced back to the original participants in the referenced clinical trials or cohort studies. This study has received IRB exemption approval from the University of North Carolina at Chapel Hill.

### Data source

Data from the placebo arm of a phase 3 clinical trial (ClinicalTrials.gov ID: NCT00409188) were obtained from Project Data Sphere.(19) This randomized, double-blind trial enrolled participants with unresectable stage III non-small-cell lung cancer (NSCLC) who had completed chemoradiotherapy within 4–12 weeks before randomization and demonstrated either stable disease or an objective response. (20) The study aimed to evaluate whether the MUC1 antigen-specific cancer immunotherapy, tecemotide, could improve overall survival. However, no significant difference in overall survival was observed between the tecemotide treatment arm and the placebo arm.(20)

### Data Exclusions and Imputations

All available participants from the placebo arm were incorporated into the analysis, including those excluded from primary clinical analyses due to a clinical hold. (20) Best supportive care provided for participants during the study may include but not limited to psychosocial support, nutrition support, and other supportive therapies. Each participants’ self-reported LCSS scores (15) throughout the study were extracted, along with baseline demographics, baseline liver and kidney function data, and OS and PFS outcomes. The patient-reported LCSS includes nine visual analogue scales (ranging from 0 – 100 millimeter) that assess the following dimensions of disease: loss of appetite, fatigue, cough, dyspnea, hemoptysis, pain, lung cancer-related symptoms, ability to perform daily activities, and QoL. Lower scores indicate fewer symptoms and better QoL. (21) In this study, LCSS scores were assessed at baseline, at weeks 2, 5, and 8, and every 6 weeks from week 13 until disease progression. Assessments continued at 6 and 12 weeks after progression and every 12 weeks thereafter. (20) For analysis, the average of the nine self-reported LCSS items at each time point was calculated, with the mean LCSS score being used as an overall measure of patient-reported symptoms and QoL. Any LCSS measurements with missing data for any of the nine items were excluded from the analyses. No imputation was performed for missing data.

### Population PRO progression model and simulation

To capture the PRO trajectories in the NSCLC population, a mathematical PRO progression model was developed and optimized using individual participants’ longitudinal LCSS scores. Although the study cohort did not receive any active treatment, many participants showed improved PROs at the beginning of the study, likely due to psychological effects. Given the subjective nature of PROs, a placebo effect was incorporated into in the model. The final model accounted for both disease progression and placebo effects on PRO dynamics. (Equation 1)

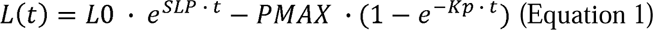

Where *L(t)* is the LCSS score changes over time, *L0* is the baseline LCSS score, *SLP* is the exponential PRO progression rate (1/day), *PMAX* is the maximum placebo effect on LCSS score, *Kp* is the placebo effect offsite rate (1/day), and *t* is the time in days.

Given the substantial variability in patient-reported LCSS scores, a population modeling approach was applied to estimate the dynamic parameters of LCSS score trajectories. This approach leveraged information from the entire analysis population, enabling accurate parameter estimation even for participants with sparse sampling. (22,23) The model was estimated using the non-linear mixed-effects (NLME) method. (24) Covariates, including population demographics and baseline clinical characteristics, were evaluated in the model to identify and describe factors that contributed to the different trajectories observed among study participants. (25) Model performance was evaluated using goodness-of-fit plots (26) and visual predictive checks (VPC) (27). To ensure robustness and assess covariate effects, the final model underwent 500 non-parametric bootstrap iterations. (28) The final model was used to simulate participants’ long-term PRO progression dynamics and assess the effects of placebo on symptom relief in the NSCLC population. (29) Details of the model development and simulation procedures are provided in the **Supplementary Methods**. (25,27–30)

### Associations between PRO progression and clinical outcomes

To evaluate the association between PRO progression and clinical outcomes, participants were grouped based on their PRO progression model parameters and their survival outcomes were compared. PFS and OS were assessed for participants with different baseline LCSS score (*L0* > median vs. *L0* ≤ median), maximum placebo effect (*PMAX* > median vs. *PMAX* ≤ median), and PRO progression rate (*SLP* > median vs. *SLP* ≤ median). Survival-related baseline characteristics were narrowed down using the Least Absolute Shrinkage and Selection Operator (LASSO) machine learning algorithm.(31) Five-fold cross-validation was applied, and prognostic clinical characteristics were selected based on the λ value within one standard error of the minimum. Cox proportional hazards models for both PFS and OS were then developed to integrate these prognostic clinical characteristics with symptom progression model parameters (*L0*, *PMAX*, and *SLP*). The contribution of each covariate to PFS and OS was compared using forest plots.

### Survival Prediction Using PRO Progression Through Machine Learning Algorithms

To predict OS using PRO progression, the study cohort was randomly divided into two sets: 80% for training and 20% for testing. An XGBoost model (32) was developed using the training cohort to predict OS based on individual PRO progression model parameters (*SLP*, *L0*, and *PMAX*). The model’s performance was validated using the testing cohort, and the Area Under the Receiver Operating Characteristic (AUC-ROC) curve was generated to assess its predictive accuracy. (33) The AUC-ROC metric ranges from 0 to 1, when 1 represents a perfect model capable of making accurate survival predictions for all subjects in the testing dataset, and 0.5 indicates a model that performs no better than random guessing.(33) The prognostic values of these PRO progression parameters were evaluated using feature importance scores within the model. The higher feature importance score indicates that the parameter contributes more to the model predictions. Packages used in the machine learning model included xgboost 2.1.3, scikit-learn 1.5.2, numpy 1.24.3, and pandas 2.0.2.

### Software and Code Availability

PRO progression model development was performed using Monolix 2021R1 (Lixoft SAS, a Simulations Plus company). PRO progression model bootstrapping was conducted using Monolix 2024R1 and model simulations were performed using Simulx 2024R1. Plots and survival analyses were performed using R 4.4.1 and RStudio (Version 2022.07.1+554). Machine learning predictive model was developed in Python 3.10.2 and the development environment is Visual Studio Code. The model code is provided in the **Supplementary Code**. The data can be accessed though Project Data Sphere. (19)

## Results

### Modeling Longitudinal PRO Progression Trajectories and Variability in NSCLC

The workflow of this study is provided in **Figure 1A**, where 6219 longitudinal, self-reported LCSS score measurements were collected from 481 individual participants receiving placebo treatment. (**Figure 1B**) Participant demographics, baseline clinical characteristics, and survival data were summarized in **Table 1** and **Table S1**. Individual participant longitudinal LCSS scores and change from baseline over time are shown in **Figure 1C** and **Figure 1D**, respectively. Significant variability in PRO progression was observed, which stems from differences in study duration and assessment times between participants, as well as within-participant fluctuations during the study due to the subjectivity of PROs and their sensitivity to individual experiences.

**Figure 1.**
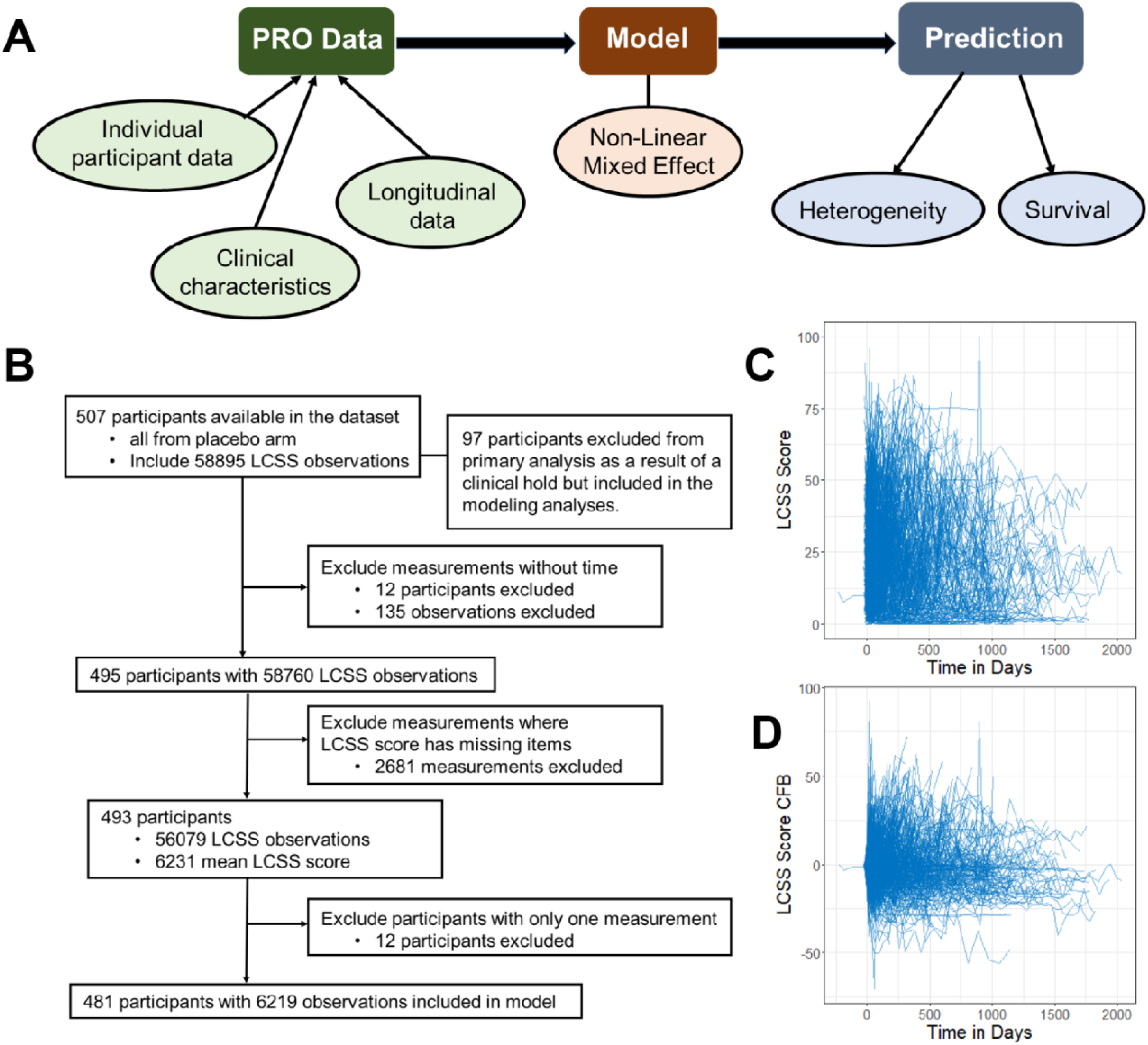
Study workflow, data inclusion and exclusion criteria, and data visualization. (A) Diagram illustrating the study workflow. (B) Diagram summarizing the inclusion and exclusion criteria for data. (C-D) Visualization of raw LCSS scores over time (C) and change from baseline in LCSS scores over time (D), with each line representing an individual participant. PRO, patient reported outcome; LCSS, Lung Cancer Symptom Scale; CFB, change from baseline.

**Table 1.** Baseline demographics, clinical characteristics and survival.

|  |  |
| --- | --- |
|  | Total (N=481) |
| Age, years | 61.0 (24.0 – 83.0) |
| Sex |  |
| Male | 327 (68.0%) |
| Female | 154 (32.0%) |
| Weight, Kg | 74.5 (38.6 – 140.8) |
| Missing | 70 (14.6%) |
| Race/Ethnicity |  |
| White | 445 (92.5%) |
| Black or African American | 5 (1.0%) |
| Asian/Pacific islander | 20 (4.2%) |
| Latino/Hispanic | 8 (1.7%) |
| Other or Missing | 3 (0.6%) |
| Type of initial chemoradiotherapy |  |
| Concurrent | 292 (60.7%) |
| Sequential | 183 (38.1%) |
| Missing | 6 (1.2%) |
| Smoking status |  |
| No | 33 (6.9%) |
| Yes | 448 (93.1%) |
| ECOG performance status |  |
| 0 | 203 (42.2%) |
| 1 | 278 (57.8%) |
| Response to initial chemoradiotherapy |  |
| Objective response | 337 (70%) |
| Stable disease | 144 (30%) |
| Overall Survival, months | 20.9 (1.7 – 65.7) |
| Missing | 93 (19.3%) |
| Progression-free survival, months | 5.9 (0.2 – 60.1) |
| Missing | 152 (31.6%) |

Data are number (%) or median (range), unless otherwise specified.
ECOG, Eastern Cooperative Oncology Group.

The PRO progression model successfully captured the dynamics of individual participants’ LCSS scores under disease progression and effectively accounted for placebo effects on PRO data. The use of a population modeling approach accommodated for data sparsity and variability by leveraging information from all participants. The model parameters are summarized in **Table S2**. **Figure 2A** presents the PRO progression profiles of nine randomly selected participants, showing the individual predictions of the model align well with the observations. **Figure 2B** shows the goodness-of-fit plots for the population PRO progression model, with no apparent biases, indicating good model performance. Additional diagnostic plots, including the normality of model residual error and model random effects distribution (**Figure S1** and **Figure S2**), further confirm the reliability of the model. The model VPC stratified by patients’ Eastern Cooperative Oncology Group (ECOG) performance status and response to initial chemotherapy are shown in **Figure 2C**. The observed median, along with the 5^th^/95^th^ percentiles, all fall within the predicted intervals, demonstrating the model performs well across all subpopulations.

**Figure 2.**
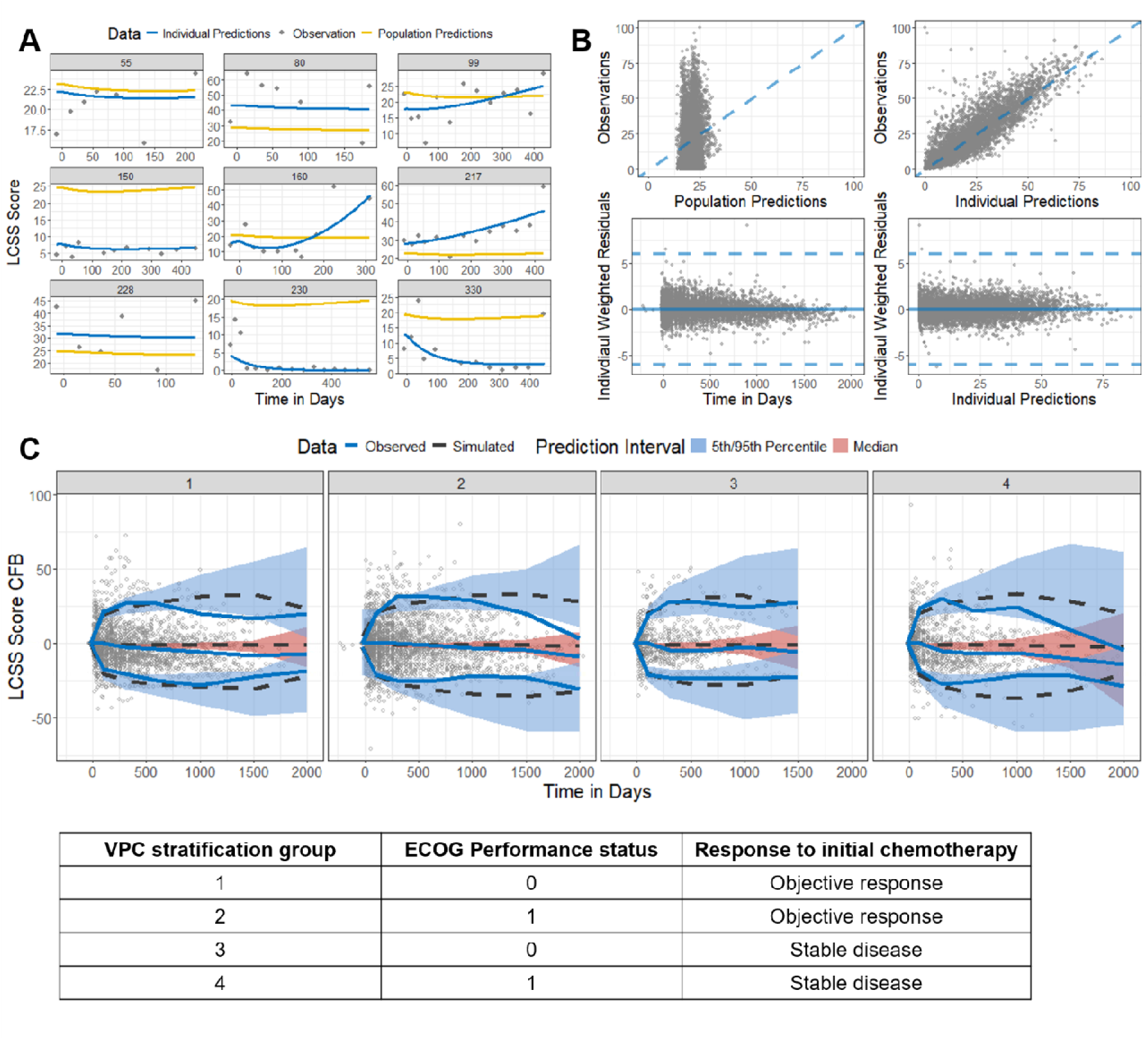
Final model captured symptom progression trajectories. (A) Plots of LCSS scores over time for nine randomly selected participants. Gray circles represent observed data, while yellow and blue solid lines represent population and individual model predictions, respectively. (B) Top left: Observations versus population predictions. Top right: observations versus individual predictions. The blue dashed lines represent the lines of identify. Bottom left: individual weighted residuals versus time (in days). Bottom right: individual weighted residuals versus individual predictions. The blue solid line represents the individual weighted residuals = 0, and dashed lines represent the individual weighted residuals ± 6. (C) VPC of LCSS scores change from baseline stratified by ECOG performance status and response to initial chemoradiotherapy. The observed data are represented by gray circles and blue solid lines (median and 5th/95th percentiles). The simulated data based on the index population (1000 simulations) are represented by the black dashed line with red shaded area (90% PI of median), or black dashed line with blue shaded area (90% PI of 5th/95th percentiles). LCSS, Lung Cancer Symptom Scale; CFB, change from baseline; VPC, visual predictive check; PI, prediction interval; ECOG, Eastern Cooperative Oncology Group.

### Covariate Analysis and Predicted PRO Progression Dynamics in NSCLC

Participants demographics (age, gender, race, ethnicity) and baseline clinical characteristics were evaluated as covariates in the model. Non-parametric bootstrap from 500 times resampling the index population was performed to assess significant covariates effect. (**Figure 3A**) ECOG performance status was identified as a significant covariate for the baseline LCSS score. Participants with a baseline ECOG performance status of “restricted in physically strenuous activity but ambulatory and able to carry out work of a light or sedentary nature” had a 28% higher baseline LCSS score compared to those with a baseline status of “fully active, able to carry on all pre-disease performance without restriction”. The type of initial chemoradiotherapy was found to significantly impact the placebo effect size. Participants who received sequential chemoradiotherapy had a 39% lower placebo effect compared to those who received concurrent chemoradiotherapy. Additionally, the response to initial chemoradiotherapy was the only significant covariate influencing PRO progression rate. Participants with stable disease after their initial chemoradiotherapy experienced a 75% faster PRO progression rate compared to those with an objective response.

**Figure 3.**
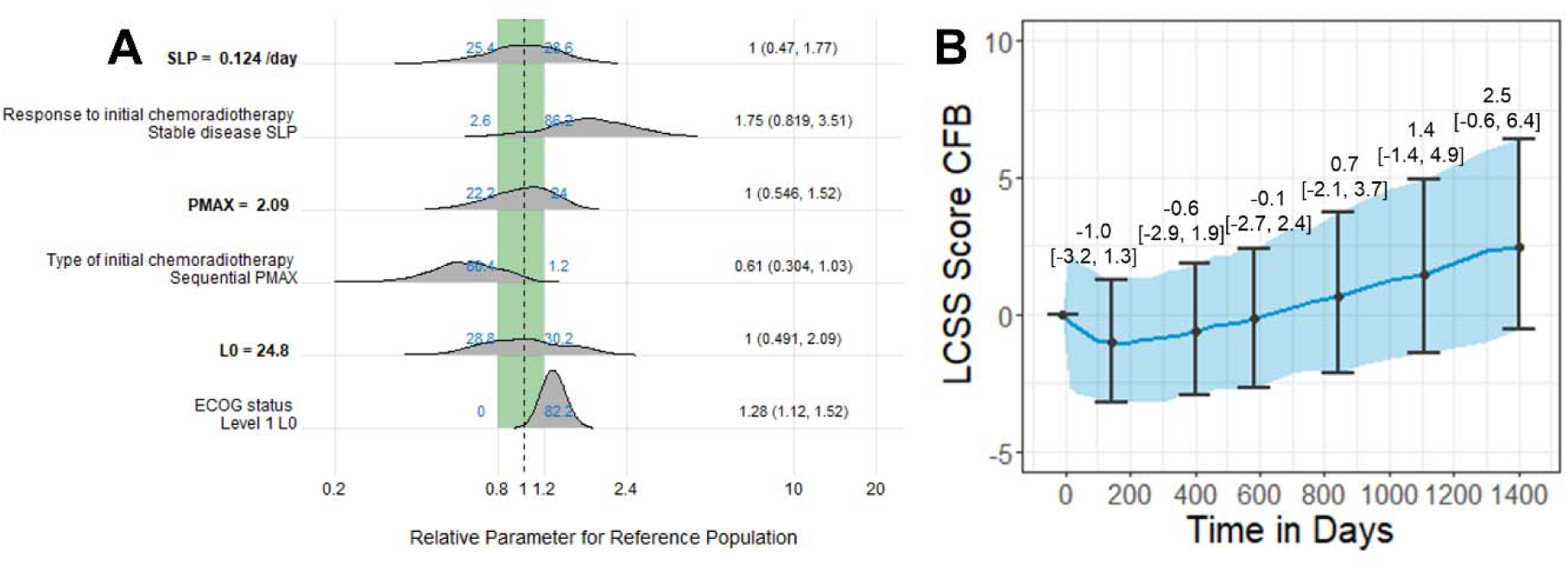
Assessment of key covariate effects on PRO progression dynamics. (A) Non-parametric bootstrap for final model was performed 500 times using sampling with replacement to evaluate the effects of key covariates. The ratio of *SLP* (PRO progression rate), *PMAX* (maximum placebo effect), and *L0* (baseline LCSS score) for each covariate estimate compared to the reference population estimate was estimated for each trial. The reference populations for *SLP*, *PMAX*, and *L0* are: participants who with an objective response to initial chemotherapy, participants who received concurrent initial chemotherapy, and participants with ECOG performance status = 0, respectively. The gray density distributions represent the ratios across all bootstrap runs, while blue numbers indicate the fractions of bootstrap runs with ratios less than 0.8 (left) or greater than 1.2 (right). Black numbers on the right y-axis show the median (5^th^ and 95^th^ percentiles) of the covariate estimate ratios. The green shaded area represents the ratio range from 0.8 to 1.2, and the black vertical dashed line indicates a ratio of 1. (B) Model-predicted LCSS scores over time due to PRO progression. Circles represent the median, and bars represent the 95% CI of the LCSS score CFB. The median and 95% CI are derived from 1000 simulations. PRO, patient-reported outcome; CI, confidence interval; LCSS, Lung Cancer Symptom Scale; ECOG, Eastern Cooperative Oncology Group.

The model was applied to predict long-term symptom progression dynamics in the NSCLC population. Model-predicted LCSS scores change from baseline over time is shown in **Figure 3B**. Heterogeneity in lung cancer populations was incorporated in the model predictions, with each sub-population equally represented in the simulations. (**Table S3**) The exponential progression pattern captured by the model indicates the rapid symptom worsening associated with advanced stages and heavy symptom burden. The maximum placebo effect in the NSCLC population was estimated as 1.0-point decrease in the LCSS score (95% confidence interval [CI], -1.3 to 3.2). The predicted time to symptom progression, measured as the model-predicted LCSS score increase from nadir, was 229 days (95% CI, 15 – 583), aligning with the primary clinical analysis estimate of 11.4 months.(20)

### Linking PRO Dynamics to Survival Outcomes Prediction in NSCLC

To explore the relationship between PRO progression and survival outcomes, sub-group analysis was conducted based on the individual PRO progression model parameters for each participant. As shown in **Figure 4A** and **4B**, OS was significantly associated with the maximum placebo effects (*PMAX*, p-value = 0.036) and the PRO progression rate (*SLP*, p-value = 0.00012). In contrast, baseline LCSS score had no significant impact on OS (p-value =0.75, **Figure 4C**). Using LASSO algorithms, five prognostic variables for OS were identified: sex, creatinine, bilirubin, aspartate aminotransferase (AST), and albumin (ALB). (**Figure S3A**) After adjusting for these survival-relevant baseline characteristics, *PMAX* and *SLP* remained significantly associated with OS. A higher PRO progression rate was linked to poorer survival (Hazard ratio [HR] 1.13, 95% CI: 1.076 – 1.18), while a greater placebo effect was associated with improved survival (HR 0.93, 95% CI: 0.883 – 0.99) (**Figure 4D**).

**Figure 4.**
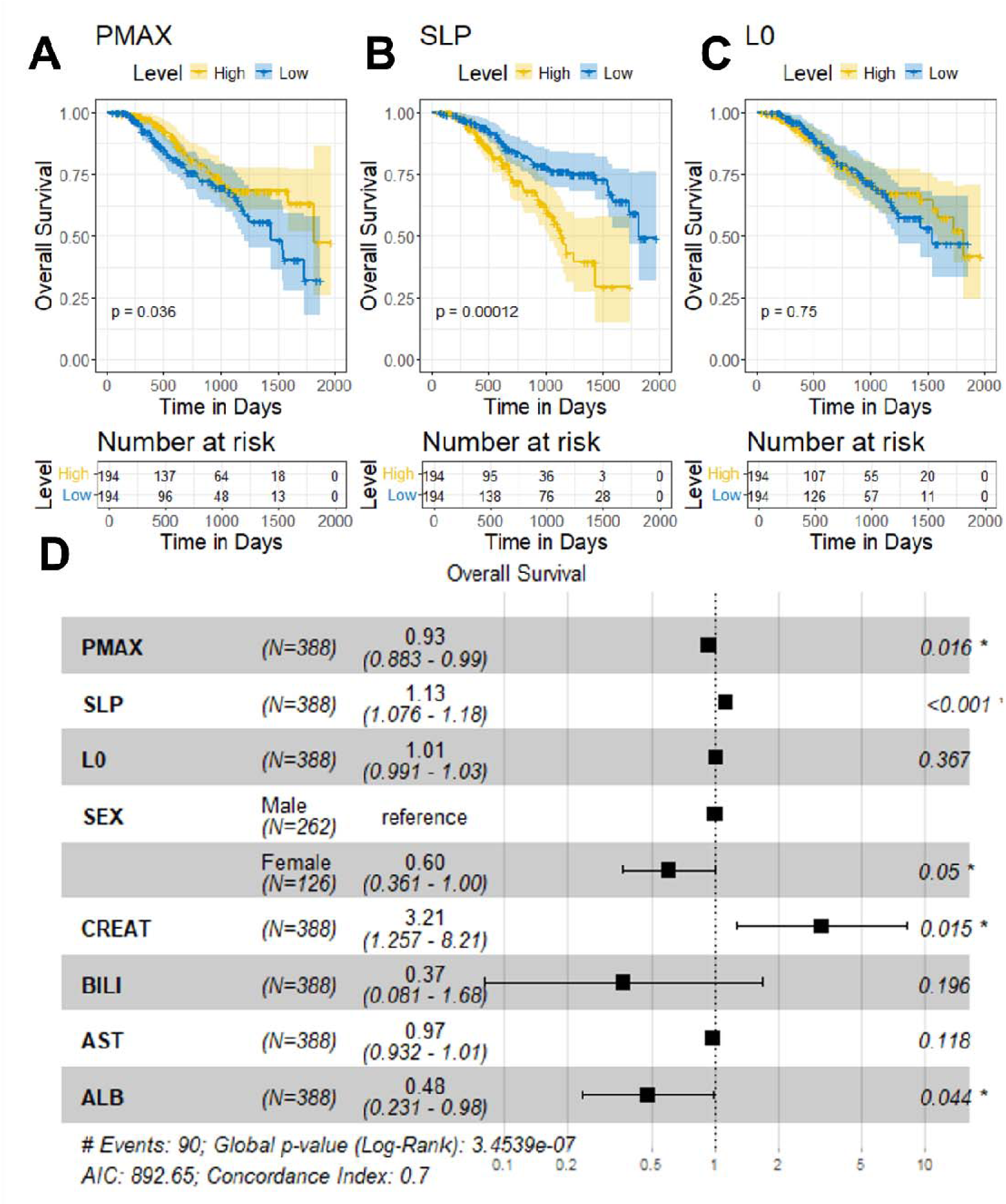
Associations between symptom progression parameters and overall survival. (A-C) Overall survival stratified by maximum placebo effects (*PMAX*), PRO progression rate (*SLP*), or baseline LCSS score (*L0*). The “High” level indicates parameters greater than the median, while the “Low” level indicates parameters less than or equal to the median. The shaded area represents the 95% CI, and p-values are calculated using the log-rank test. (D) Cox proportional hazards model incorporating model parameters and LASSO-selected survival-related clinical characteristics. The black boxes with horizontal error bars represent hazard ratio estimates with 95% CI. P-values for each covariate are labeled on the right. PRO, patient-reported outcome; LCSS, Lung Cancer Symptom Scale; LASSO, least absolute shrinkage and selection operator; CI, confidence interval; CREAT, creatinine; BILI, bilirubin; AST, aspartate aminotransferase; ALB, albumin.

Similar analyses were performed for PFS. Sex, neutrophil count, AST, and smoking status were identified as prognostic factors for PFS by LASSO algorithm. (**Figure S3B**) The symptom progression model parameters showed no strong correlations with PFS when stratified by median values (**Figure S4A-C**). However, after adjusting for PFS-related baseline characteristics identified by LASSO, *PMAX* and *SLP* demonstrated statistically significantly associations with PFS. A higher PRO progression rate was linked to worse PFS (HR 1.06, 95% CI: 1.04 – 1.1), while a greater placebo effect was associated with better PFS (HR 0.98, 95% CI: 0.96 – 1.0) (**Figure S4D**).

### Predicting Overall Survival Through PRO Progression Trajectories

An XGBoost machine learning model was developed to predict survival in NSCLC patients using individual PRO progression model parameters (*L0*, *SLP*, and *PMAX*). The model achieved an AUC-ROC of 0.78, indicating a 78% probability of correctly distinguishing between survive and death cases. (**Figure S5**) This level of discrimination (0.7 – 0.8) is considered acceptable given the limited sample size used for model training. Among the three parameters, *SLP* was the most influential, with a feature importance score of 0.345, followed by *L0* and *PMAX*, with scores of 0.33 and 0.324, respectively. These findings suggest that PRO progression trajectories can predict overall survival and may serve as complementary surrogate endpoints in clinical decision-making.

## Discussion

PROs are patient-centered, non-invasive tools for monitoring symptoms, functioning, and QoL in clinical trials and routine care.(2) With advancements in digital health, PROs can now be collected remotely, allowing for regular assessments without requiring clinic visits.(34,35) This enhances their accessibility and generalizability compared to traditional cancer evaluation methods, such as tumor volume. Despite these advantages, PROs have been underutilized in clinical decision-making due to the unclear correlation between PROs and survival outcomes.

In this study, long-term PRO progression in NSCLC participants was analyzed using patient-reported LCSS data and explored the associations between PRO progression dynamics and survival outcomes. While the large variability in PRO data would typically present significant challenges to analysis, this was addressed through the application of a population modeling approach that integrated individual participant longitudinal LCSS score data. The model effectively described long-term PRO progression trajectories, and it was found that PRO dynamics are prognostic of survival outcomes in NSCLC patients. In summary, a robust workflow was developed to analyze PRO progression and predict OS using model-informed PRO trajectories. This workflow has the potential to be applied to other PRO clinical endpoints, enhancing their utility in clinical decision-making and improving patient-centered care.

During the development of the PRO progression model, both linear and exponential models were evaluated. The exponential model provided a better fit to the data. Those who achieved an objective response to prior chemoradiotherapy exhibited slower PRO progression than those who had stable disease response, even following the completion of chemoradiotherapy. The exponential model effectively captures the slow initial progression of PROs observed at the start of treatment and the rapid escalation of symptom burden in late-stage disease, reflecting the accelerated progression of symptoms as the disease advances. This aligns with the observed clinical trajectory of the disease. (36)

We further evaluated the association between participants’ PRO progression dynamics and survival outcomes. The PRO progression rate was significantly associated with both OS and PFS, suggesting that PRO dynamics are linked to survival outcomes. In contrast, the baseline LCSS score was not associated with either OS or PFS, indicating that it is not the absolute value of PROs, but rather their dynamics, that are meaningful. This finding is consistent with prior studies demonstrating that dynamic changes in symptoms, such as pain, fatigue, and dyspnea, often predict survival outcomes in lung and other cancers more effectively than single-time point scores. (10)

Finally, the model-estimated PRO parameters were used to predict OS, achieving acceptable performance with a machine learning approach. This underscores the potential of PRO dynamics as complementary surrogate endpoints alongside PFS to inform clinical decision-making. However, the predictive performance is constrained by the limited sample size of the analysis dataset, highlighting the need for further refinement. Future research will focus on scaling this PRO progression model workflow to larger datasets, enabling the development of more robust predictive models and potentially enhancing the accuracy and utility of PRO-based survival predictions in clinical practice.

While most analyses linking PRO data to survival have used landmark analyses (9,37), our approach incorporated longitudinal data from all visits, making it less sensitive to missed or delayed assessments. One application of this approach in patients with NSCLC is to assess the probability of changes in PROs being transient versus indicative of underlying disease progression. Clinicians could use this information to determine when intervention is necessary. There is precedent for using population analyses in adaptive dosing, where time-varying disease aspects (e.g., progression trajectory) inform dose adjustments (i.e., clinical intervention). (22,38) Similarly, this analysis could optimize oncology clinical trials by evaluating protocol-specified visit schedules to ensure visits are frequent enough to capture PRO changes and disease progression effectively.

Our study has some limitations. Data from the treatment arm of this clinical trial were not available for analysis, preventing assessment of the impact of treatment on PRO progression in NSCLC participants. This lack of data brings additional challenges for model validation. A recent perspective paper has called for stakeholders to share individual participant data from oncology clinical trials for scientific purposes. (39) Such initiatives could pave the way for a deeper understanding of not only the clinical questions discussed here, but cancer disease progression more broadly. Another limitation of this study was that the average of all nine LCSS items was used to represent the overall LCSS status for patients with NSCLC. While this approach simplifies computation and interpretation, it is somewhat arbitrary and may result in a loss of information at the individual item level. By combining the present approach with the statistical framework of item-response theory (IRT), the analysis and interpretation of responses could be expanded to each individual question. This additional complexity would allow for use of LCSS measurements where responses to some questions were missing, as well as accounting for questions with differing sensitivity to changes in disease and which may be assessing different aspects of disease that may progress independently. (40)

In summary, this study developed a population modeling platform to analyze PRO progression in NSCLC participants using longitudinal LCSS data. The model can predict long-term symptom progression while accommodating the substantial variability in PRO data. The PRO progression parameters derived from the model were shown to be prognostic of clinical outcomes. This framework is generalizable to other PRO datasets, integrating individual participant data to provide insights into PRO dynamics for each participant. It holds significant promise for enhancing clinical decision-making and optimizing the design and evaluation of clinical trials.

## Data Availability

The data can be accessed though Project Data Sphere.

https://data.projectdatasphere.org/projectdatasphere/html/home

