## Supplementary Materials for "Leveraging Longitudinal Patient-Reported Outcomes Trajectories to Predict Survival in Non-Small-Cell Lung Cancer"

#### Supplementary Methods

##### 1. Model development

Model development used Monolix 2021R1 (Lixoft SAS, a Simulations Plus company). Population parameter estimation used stochastic approximation expectation-maximization (SAEM) algorithm and individual parameters were obtained from conditional modes.

##### 1.1. Bounded data transformation

For LCSS scores that are at the boundaries (0 or 100), 0 was replaced with 0.05 and 100 was replaced with 99.95 to avoid error caused by the bounded data. As the fraction of the data at boundaries are less than 5% this was not anticipated to have a significant impact on analysis.

##### 1.2 Base model development

An asymptotic exponential function with time was used to account for the placebo effect on Lung Cancer Symptom Scale (LCSS) score in participants:

$Pbo(t) = PMAX \cdot (1 - e^{-Kp \cdot t})$ (Equation 1)

Where *Pbo(t)* is the LCSS scores change over time due to the placebo effects, *PMAX* is the maximum placebo effects, *Kp* is the placebo effect offsite rate (1/day).

Both linear and exponential function with time accounted for PRO progression were tested and the exponential function model was selected as the final model due to better performance:

$Dis(t) = L0 \cdot e^{SLP \cdot t}$ (Equation 2)

Where *Dis(t)* is the LCSS scores change over time due to the PRO progression, *L0* is the baseline LCSS score, *SLP* is the PRO progression rate (1/day).

The LCSS score dynamics *L(t)* were described as the combination effects of placebo *Pbo(t)* and PRO progression *Dis(t)*:

$L(t) = Dis(t) - Pbo(t)$ (Equation 3)

**1.3 Random effects model development**

Inter-individual variability (IIV) was assumed to be log-normally distributed for *PMAX* and *SLP* parameters:

$P_i = \theta_P \cdot e^{\eta_i}$ (Equation 4)

Where $P_i$ is the individual value for parameter, *P*, in the $i^{th}$ subject, and $\theta_P$ is the population typical value for parameter *P*, and $\eta$ is an independent random variable describing the variability in *P* among subjects with a mean of 0 and variance $\omega^2$.

Both log-normally distributed and normally distributed random effects were tested on parameter *L0* but were not able to describe the shape of parameter. Therefore, box-cox transformation(1) was performed on *L0*:

$L0_i = \theta_{L0} \cdot e^{\frac{(e^{\lambda \cdot \eta\_L0_i}) - 1}{\lambda}}$ (Equation 5)

Where $L0_i$ is the individual value for parameter *L0*, in the $i^{th}$ subject, and $\theta_{L0}$ is the population typical value for parameter *L0*, and $\eta\_L0$ is an independent random variable describing the variability in *L0* among subjects, and λ is the shape parameter that transform a normal distribution to a left or right skewed distribution.

A logit transformation with bounds (0 to 100) was applied in the residual error model. Both additive and proportional residual error models were used to describe the unexplained variability:

$\log(L_{ij}/(100 - L_{ij})) = \log(IPRED_{ij}/(100 - IPRED_{ij})) \cdot (1 + \varepsilon_{ij,b}) + \varepsilon_{ij,a}$ (Equation 6)

Where $L_{ij}$ is the observed LCSS score in subject *i* at time *j*. $IPRED_{ij}$ is the model predicted LCSS score for subject *i* at time *j*. $\varepsilon_{ij,a}$ is the additive error term and $\varepsilon_{ij,b}$ is the proportional error term, both with mean of 0 and variance of $\sigma^2$.

**1.4 Covariate model development**

Based on exploratory analysis and clinical relevance, potential baseline covariates were evaluated on parameters *L0*, *SLP*, and *PMAX*, including age, gender, race, ethnicity, height, body weight, type of initial chemoradiotherapy (concurrent or sequential), leukocytes (WBC), neutrophils (NEUT), creatinine (CREAT), Alanine Aminotransferase (ALT), bilirubin (BILI), Alkaline Phosphatase (ALP), Aspartate Aminotransferase (AST), Albumin (ALB), previous depression or anxiety history, smoking status, Eastern Cooperative Oncology Group (ECOG) performance status, response to initial chemoradiotherapy (objective response or stable disease). These covariates were initially plotted against individual parameters to identify any relationships. If a trend was observed, the covariate was included in the full model.

Full fixed effect modeling approach was used for covariate model development.(2) Covariates that show a trend when plotted against individual parameters were added to the base model simultaneously. Correlations between covariates were assessed, and in cases where strong correlations existed, the covariate that was more clinically relevant or with greater statistical significance was selected to be included.

The effect of a categorical covariate on a parameter was represented as a discrete relationship. For example, the effect of ECOG performance status on a parameter *P* was described as exponential effect:

$$P_i = \theta_P \cdot e^{Cov_{ECOG}} \text{ for } Cov_{ECOG} = \begin{cases} 0 & if\ ECOG\ status = 0 \\ \theta_{ECOG} & if\ ECOG\ status = 1 \end{cases} \text{(Equation 7)}$$

Where $\theta_{ECOG}$ is the estimable parameter for the effect of ECOG status 1 on parameter *P*.

The effect of a continuous covariate on a parameter was presented as exponential relationship. For example, the effect of ALB on a parameter $P$ was described as:

$P_i = \theta_P \cdot e^{\theta_{ALB} \cdot ALB}$ (Equation 8)

Where $\theta_{ALB}$ is the estimable parameter for the effect of ALB on parameter $P$.

Missing values for covariates were imputed with the population median (for continuous covariates) or mode (for categorical covariates) during the covariate analyses.

#### 2. Assessment of Model Performance

##### 2.1 Goodness of Fit

Model goodness-of-fit was assessed by changes in the minimum objective function value (OFV). Diagnostic plots used to assess model performance included:

- Observations versus population predictions or individual predictions.
- Individual weighted residuals versus time or population predictions.
- Normality of individual weighted residuals distributions by evaluation of distribution density and quantile-quantile plots.
- Normality of random effect distributions by evaluation of distribution density plots.
- Individual predicted LCSS scores over time profiles overlaid with observations.

##### 2.2 VPC checks

The predictive performance of the final model was evaluated by visual predictive check (VPC)(3) based on 1000 simulations of the index dataset. Subpopulations with differing ECOG performance status (0 or 1) and response to initial chemoradiotherapy (objective response or stable disease) were used to derive summaries of the predictive performance of LCSS score change from baseline (CFB) dynamics.

#### 3. Bootstrap

Non-parametric bootstrap was performed to evaluate model robustness and covariates effects. Bootstrap resampling was performed 500 times using sampling with replacement.(4) Population parameters were estimated for each bootstrap dataset, with the median and 95% confidence intervals for each parameter calculated to assess parameter uncertainty. The bootstrap was performed using Monolix 2024R1.

**4. Model simulations**

The final model was used to perform simulations for 1000 studies. Each study was simulated with a new set of parameters from final model typical estimates and the covariance-variance matrix (parameter with uncertainty).(5) In each study, 8 subpopulations with different covariates were simulated. (**Table S3**) The simulation results were used to predict the LCSS score dynamics and CFB dynamics after 1400 days. The simulations were performed using Simulx 2024R1 (Lixoft SAS, a Simulations Plus company).

**Supplementary Tables and Figures**

**Table S1. Baseline clinical characteristics additional to Table 1.**

| | Total (N=481) |
|---|---|
| Height, cm | 171 (144 – 194) |
| Missing | 68 (14.1%) |
| Alanine Aminotransferase, IU/L | 17 (5 – 112) |
| Missing | 67 (13.9%) |
| Bilirubin, mg/dL | 0.3 (0.1 – 1.3) |
| Missing | 68 (14.1%) |
| Alkaline Phosphatase, IU/L | 78 (32 – 362) |
| Missing | 69 (14.3%) |
| Aspartate Aminotransferase, IU/L | 19.5 (7 – 117) |
| Missing | 67 (13.9) |
| Albumin, g/dL | 4.2 (2.6 – 5.2) |
| Missing | 66 (13.7%) |
| Prior depression/anxiety history | |
| No | 442 (91.9%) |
| Yes | 39 (8.1%) |
| Leukocytes, ×10E3/µl | 5.7 (2.8 – 16.6) |
| Missing | 67 (13.9%) |
| Neutrophils, ×10E3/µl | 4.14 (1.77 – 13.28) |
| Missing | 75 (15.6%) |
| Creatinine, mg/dL | 0.9 (0.5 – 1.9) |
| Missing | 67 (13.9%) |

Data are number (%) or median (range), unless otherwise specified.

**Table S2. Final model parameter estimates.**

| Parameter | Value | RSE (%) | Bootstrap median | Bootstrap 95% CI | SHR (%) |
|---|---|---|---|---|---|
| **Population parameters** | | | | | |
| Baseline LCSS score ($\theta_{L0}$) | 37.1 | 9.7 | 24.8 | 12.0, 52.2 | |
| Box-Cox transformed variability ($\eta\_L0$) | 0 Fixed | | 0 Fixed | | |
| Box-Cox transformed parameter (λ) | -0.892 | 1.03 | -0.581 | -0.739, -0.459 | |
| Maximum placebo effect (PMAX) | 2.48 | 12.4 | 2.09 | 1.126, 3.211 | |
| Placebo effect offset rate (Kp, 1/day) | 0.0116 | 4.75 | 0.01 | 0.006, 0.016 | |
| PRO progression rate, (SLP, 1/day) | $0.143\times10^{-3}$ | 19.2 | $0.124\times10^{-3}$ | $0.058\times10^{-3}$, $0.22\times10^{-3}$ | |
| Albumin covariate effect on baseline ($\theta_{ALB\_L0}$) | -0.18 | 11.6 | -0.187 | -0.369, 0.0002 | |
| Leukocytes covariate effect on baseline ($\theta_{WBC\_L0}$) | 0.012 | 19.5 | 0.045 | 0.002, 0.092 | |
| ECOG performance status 1 covariate effect on baseline ($\theta_{ECOG\_L0}$) | 0.211 | 4.81 | 0.249 | 0.107, 0.419 | |
| Initial chemotherapy type (Sequential) covariate effect on | -0.379 | 53.8 | -0.494 | -1.191, 0.035 | |

| | | | | | |
|---|---|---|---|---|---|
| maximum placebo effect ($\theta_{PRCANCER\_PMAX}$) | | | | | |
| Response to initial chemoradiotherapy (stable disease) covariate effect on PRO progression rate ($\theta_{BRESPONSE\_SLP}$) | 0.439 | 71.3 | 0.562 | -0.228, 1.258 | |
| **Inter-individual variability (standard deviation)** | | | | | |
| Standard deviation of random effects on Box-Cox transformed baseline ($\Omega_{L0}$) | 0.775 | 3.86 | 0.858 | 0.744, 0.955 | 0.51 |
| Standard deviation of random effects on maximum placebo effect ($\Omega_{PMAX}$) | 1.16 | 6.5 | 1.278 | 0.986, 1.662 | 0.362 |
| Standard deviation of random effects on PRO progression rate ($\Omega_{SLP}$) | 2.14 | 5.46 | 2.315 | 1.971, 2.747 | 3.55 |
| **Residual unexplained variability** | | | | | |
| Additive residual error ($\boldsymbol{a}$) | 0.341 | 2.46 | 0.325 | 0.287, 0.367 | |
| Proportional residual error ($\boldsymbol{b}$) | 0.186 | 2.9 | 0.192 | 0.154, 0.224 | |

RSE, relative standard error; CI, confidence interval; SHR, shrinkage.

**Table S3. Subpopulation features in PRO progression dynamics simulations.**

| **Subpopulation** | **ECOG Performance status** | **Response to initial chemoradiotherapy** | **Type of initial chemoradiotherapy** |
|---|---|---|---|
| #1 | 0 | Objective response | Sequential |
| #2 | 1 | Objective response | Sequential |
| #3 | 0 | Stable disease | Sequential |
| #4 | 1 | Stable disease | Sequential |
| #5 | 0 | Objective response | Concurrent |
| #6 | 1 | Objective response | Concurrent |
| #7 | 0 | Stable disease | Concurrent |
| #8 | 1 | Stable disease | Concurrent |

### Figure S1

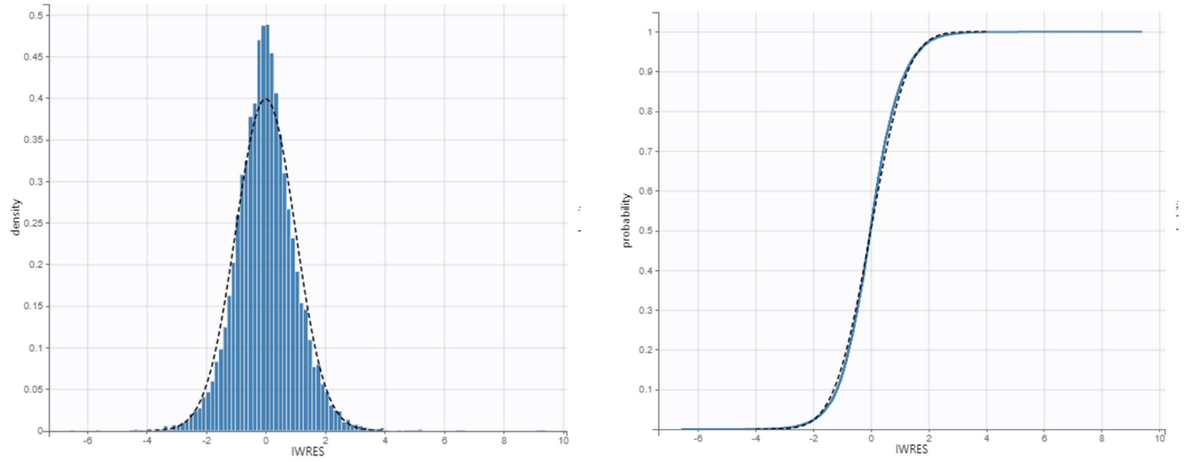


**Figure S1. Normality of IWRES distributions checks.** ***Left****:* The IWRES density (blue bars) overlay with normal distribution (black dashed line). *Right*: Probability of IWRES (blue solid line) overlay with theoretical distribution (black dashed line). The IWRES distribution overlay well with theoretical distribution indicating a good normality of residual error model. IWRES, individual weight residuals.

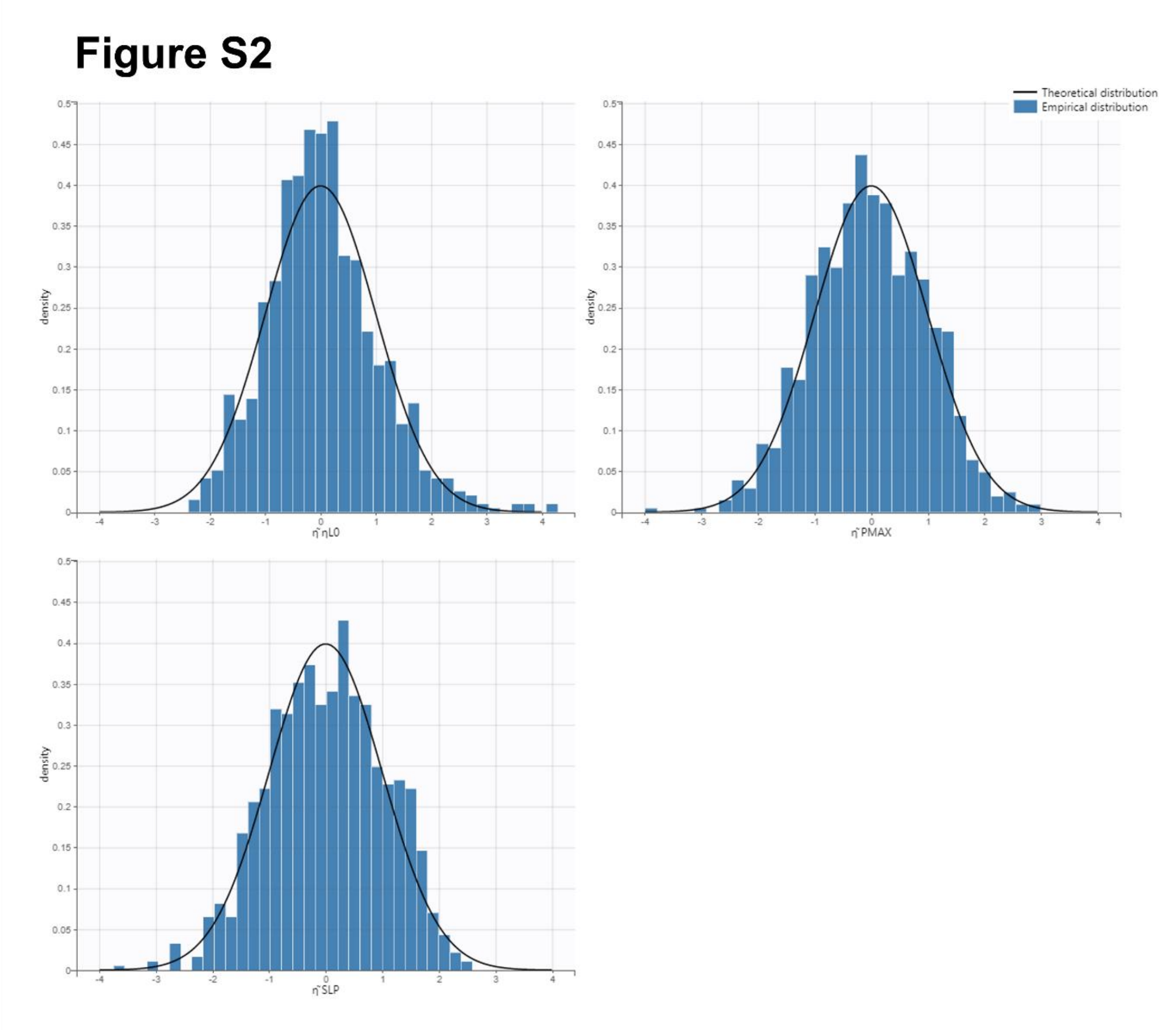


**Figure S2. Normality of η distributions checks.** The blue bars represent the distribution of model estimated random effects ηs for three parameters *L0*, *PMAX*, and *SLP*. The black solid lines represent the theoretical normal distributions. The η distributions aligned well with normal distribution indicating the appropriate selection of random effect models.

**Figure S3**

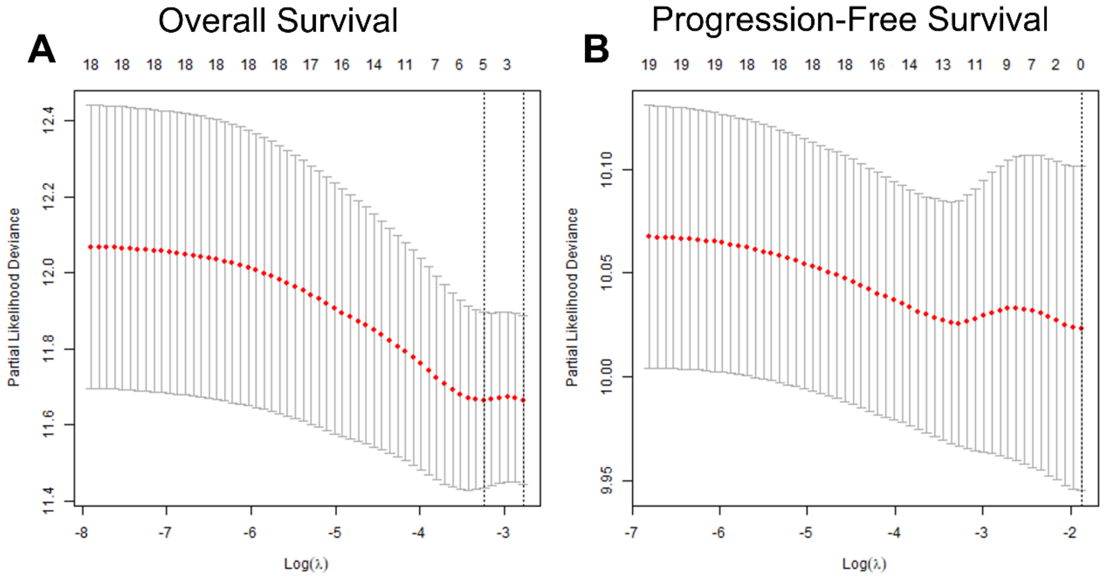


**Figure S3. The relationship between partial likelihood deviance and lambda from LASSO algorithms for overall survival (A) and progression-free survival (B).** The red dotted line represents the cross-validation curve, and the gray error bars represent the upper and lower standard deviation curves along the lambda sequence. The selected lambdas (lambda min or lambda se) are indicated by the vertical dashed lines. For overall survival related variable selection, λ = lamda.se were used to select covariates. For progression-free survival, λ = 0.01 was used to select covariates because no lamda.se was available in the cross-validation.(6)

### Figure S4

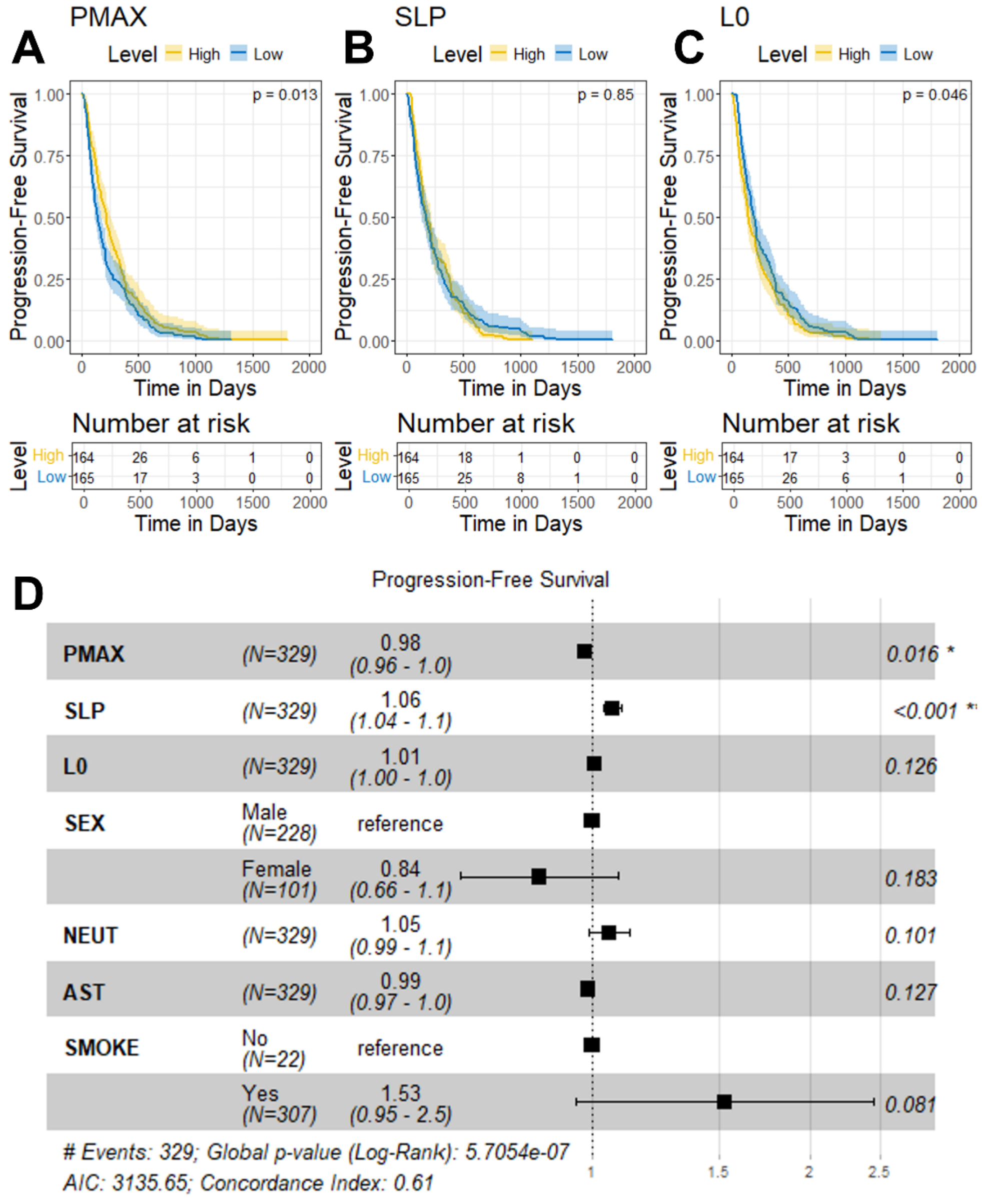


**Figure S4. Associations between symptom progression parameters and progression-free survival.** (A-C) Progression-free survival stratified by maximum placebo effects (*PMAX*), PRO progression rate (*SLP*), or baseline LCSS score (*L0*). The "High" level indicates parameters greater than the median, while the "Low" level indicates parameters less than or equal to the median. The shaded area represents the 95% CI, and p-values are calculated using the log-rank test. (D) Cox proportional hazards model incorporating model parameters and LASSO-selected survival-related clinical characteristics. The black boxes with horizontal error bars represent hazard ratio estimates with 95% CI. P-values for each covariate are labeled on the right. LCSS, Lung Cancer Symptom Scale; LASSO, least absolute shrinkage and selection operator; CI, confidence interval; NEUT, neutrophils; AST, aspartate aminotransferase; SMOKE, smoking status.

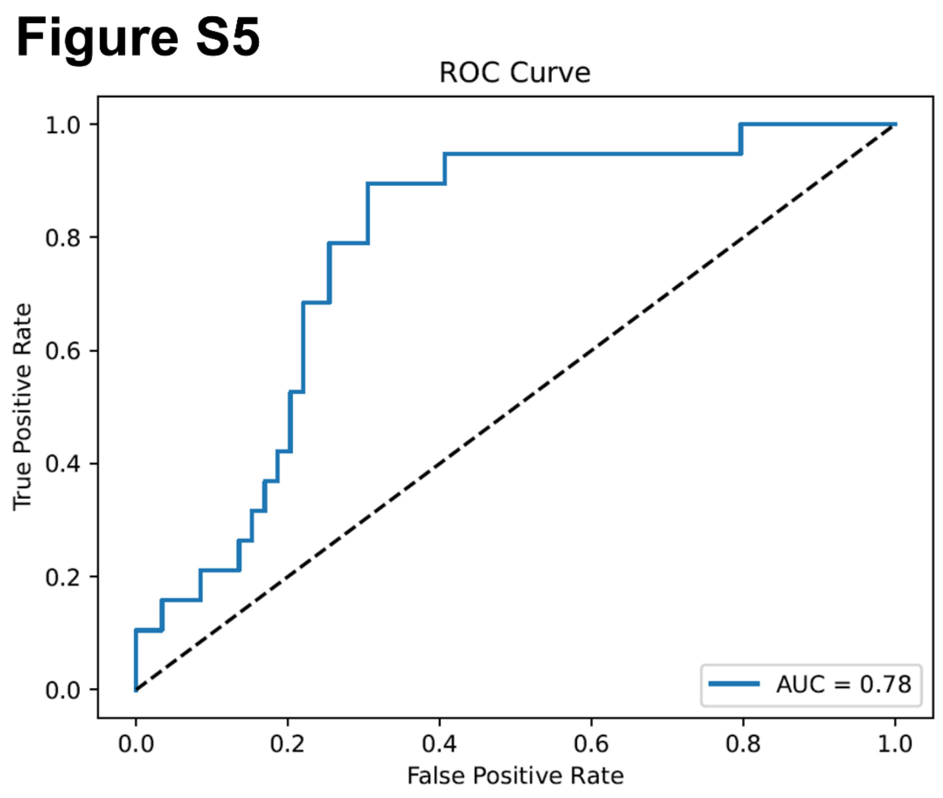


**Figure S5. AUC-ROC curve of machine learning predictive model based on symptom progression parameters.** AUC-ROC, Area Under the Receiver Operating Characteristic curve

#### Supplementary Codes

##### 1. Symptom progression model codes

```
DESCRIPTION:
 The PD model is a linear model with a linear baseline.
 The placebo effects is added.
 Disease progression is assumed.
 Box-Cox transformation is applied for L0.

[LONGITUDINAL]
 input = {L0_POP, eta_L0, lambda, PMAX, Kp, SLP, FIRSTTIME}
 FIRSTTIME     = {use=regressor}

EQUATION:
 ; Box-Cox transformation for L0
 etabar = (exp(lambda * eta_L0) -1)/lambda
 L0 = L0_POP * exp(etabar)

 ; Placebo effects
 if t < 0
   PBO = 0
 else
   PBO = PMAX * (1 - exp(-Kp * t))
 end

 ; Disease progression effect
 SLP2 = SLP * 0.001
 DIS = L0 * exp(SLP2 * (t - FIRSTTIME))
 L = DIS - PBO

 OUTPUT:
 output = {L}
 table = {L0, SLP2}
```

#### 2. Monolix settings

```
<DATAFILE>

[FILEINFO]
file='Merck_Lung_9188_LCSS_Monolix_NAreplace_13OCT2024.csv'
delimiter = comma
header = {C, USUBJID, SUBJID, AGE, SEX, RACE, ETHNIC, ARM, PRCANCER, WBC, NEUT, CREAT, ALT, BILI, 
ALP, AST, ALB, MHDEPRESSION, MHDANXIETY, SUOCCUR, HEIGHT, BSA, WEIGHT, XPSTRESN, DTHDY, DTH, PFSDY, 
PFS, BRESPONSE, B_EQ5D1, B_EQ5D2, B_EQ5D3, B_EQ5D4, B_EQ5D5, B_EQ5D6, B_LCSSO1, B_LCSSO2, B_LCSSO3, 
B_LCSSO4, B_LCSSO5, B_LCSSO6, B_LCSSS1, B_LCSSS2, B_LCSSS3, B_LCSSS4, B_LCSSS5, B_LCSSS6, B_LCSSS7, 
B_LCSSS8, B_LCSSS9, TIME, TAFR, totalLCSS, meanLCSS, FIRSTTIME, LASTTIME, LASTTIMEC, DV}

[CONTENT]
USUBJID = {use=identifier}
AGE = {use=covariate, type=continuous}
SEX = {use=covariate, type=categorical}
RACE = {use=covariate, type=categorical}
ETHNIC = {use=covariate, type=categorical}
PRCANCER = {use=covariate, type=categorical}
WBC = {use=covariate, type=continuous}
NEUT = {use=covariate, type=continuous}
CREAT = {use=covariate, type=continuous}
ALT = {use=covariate, type=continuous}
BILI = {use=covariate, type=continuous}
ALP = {use=covariate, type=continuous}
AST = {use=covariate, type=continuous}
ALB = {use=covariate, type=continuous}
MHDEPRESSION = {use=covariate, type=categorical}
MHDANXIETY = {use=covariate, type=categorical}
SUOCCUR = {use=covariate, type=categorical}
HEIGHT = {use=covariate, type=continuous}
WEIGHT = {use=covariate, type=continuous}
XPSTRESN = {use=covariate, type=categorical}
BRESPONSE = {use=covariate, type=categorical}
TIME = {use=time}
FIRSTTIME = {use=regressor}
DV = {use=observation, name=DV, type=continuous}

<MODEL>

[COVARIATE]
input = {AGE, ALB, ALP, ALT, AST, BILI, CREAT, HEIGHT, NEUT, WBC, WEIGHT, BRESPONSE, ETHNIC, 
MHDANXIETY, MHDEPRESSION, PRCANCER, RACE, SEX, SUOCCUR, XPSTRESN}

BRESPONSE = {type=categorical, categories={1, 2}}
ETHNIC = {type=categorical, categories={0, 1}}
MHDANXIETY = {type=categorical, categories={0, 1}}
MHDEPRESSION = {type=categorical, categories={0, 1}}
PRCANCER = {type=categorical, categories={1, 2}}
RACE = {type=categorical, categories={1, 2, 3, 4, 5}}
SEX = {type=categorical, categories={0, 1}}
SUOCCUR = {type=categorical, categories={0, 1}}
XPSTRESN = {type=categorical, categories={0, 1}}

[INDIVIDUAL]
input = {Kp_pop, L0_POP_pop, PMAX_pop, omega_PMAX, SLP_pop, omega_SLP, eta_L0_pop, omega_eta_L0, 
lambda_pop, ALB, beta_L0_POP_ALB, XPSTRESN, beta_L0_POP_XPSTRESN_1, WBC, beta_L0_POP_WBC, PRCANCER, 
beta_PMAX_PRCANCER_2, BRESPONSE, beta_SLP_BRESPONSE_2}
```

```
XPSTRESN = {type=categorical, categories={0, 1}}
PRCANCER = {type=categorical, categories={1, 2}}
BRESPONSE = {type=categorical, categories={1, 2}}

DEFINITION:
Kp = {distribution=logNormal, typical=Kp_pop, no-variability}
L0_POP = {distribution=logNormal, typical=L0_POP_pop, covariate={ALB, XPSTRESN, WBC},
coefficient={beta_L0_POP_ALB, {0, beta_L0_POP_XPSTRESN_1}, beta_L0_POP_WBC}, no-variability}
PMAX = {distribution=logNormal, typical=PMAX_pop, covariate=PRCANCER, coefficient={0,
beta_PMAX_PRCANCER_2}, sd=omega_PMAX}
SLP = {distribution=logNormal, typical=SLP_pop, covariate=BRESPONSE, coefficient={0,
beta_SLP_BRESPONSE_2, sd=omega_SLP}
eta_L0 = {distribution=normal, typical=eta_L0_pop, sd=omega_eta_L0}
lambda = {distribution=normal, typical=lambda_pop, no-variability}

[LONGITUDINAL]
input = {a, b}

file = 'test18_exp_PBO.txt'

DEFINITION:
DV = {distribution=logitNormal, min=0, max=100, prediction=L, errorModel=combined1(a, b)}

<FIT>
data = DV
model = DV

<PARAMETER>
Kp_pop = {value=0.01, method=MLE}
L0_POP_pop = {value=20, method=MLE}
PMAX_pop = {value=5, method=MLE}
SLP_pop = {value=1, method=MLE}
a = {value=1, method=MLE}
b = {value=0.3, method=MLE}
beta_L0_POP_ALB = {value=0, method=MLE}
beta_L0_POP_WBC = {value=0, method=MLE}
beta_L0_POP_XPSTRESN_1 = {value=0, method=MLE}
beta_PMAX_PRCANCER_2 = {value=0, method=MLE}
beta_SLP_BRESPONSE_2 = {value=0, method=MLE}
c = {value=1, method=FIXED}
eta_L0_pop = {value=0, method=FIXED}
lambda_pop = {value=-1, method=MLE}
omega_PMAX = {value=1, method=MLE}
omega_SLP = {value=1, method=MLE}
omega_eta_L0 = {value=1, method=MLE}

<MONOLIX>

[TASKS]
populationParameters()
individualParameters(method = {conditionalMean, conditionalMode })
fim(method = StochasticApproximation)
logLikelihood(method = ImportanceSampling)
plotResult(method = {indfits, parameterdistribution, covariancemodeldiagnosis,
covariatemodeldiagnosis, obspred, vpc, residualsscatter, residualsdistribution, randomeffects,
saemresults })

[SETTINGS]
GLOBAL:
exportpath = 'test18_final'
```

#### 3. Machine Learning Model Codes

```
import pandas as pd
import numpy as np
from sklearn.model_selection import train_test_split
from sklearn.metrics import roc_auc_score, roc_curve
import xgboost as xgb
import matplotlib.pyplot as plt

os_train = pd.read_csv('./OS_train.csv')
os_test = pd.read_csv('./OS_test.csv')


# Define features and target
features = ['L0_mode', 'SLP2_mode', 'PMAX_mode']
target = 'DTH'

# Prepare the training data
X_train = os_train[features]
y_train = os_train[target]

# Prepare the testing data
X_test = os_test[features]
y_test = os_test[target]


# Initialize and train the XGBoost classifier
model = xgb.XGBClassifier(objective='binary:logistic', use_label_encoder=False,
eval_metric='logloss', n_estimator=1200,max_depth=4)
model.fit(X_train, y_train)

# Make predictions on the test set
y_pred_proba = model.predict_proba(X_test)[:, 1]

# Calculate AUC
auc = roc_auc_score(y_test, y_pred_proba)
print(f"AUC: {auc}")

#Get feature importances
feature_importances = model.feature_importances_

# Print feature importance with feature names
for importance, feature_name in sorted(zip(feature_importances, features), reverse=True):
  print(f"Feature: {feature_name}, Importance: {importance}")

# Plot AUC curve
fpr, tpr, thresholds = roc_curve(y_test, y_pred_proba)
plt.plot(fpr, tpr, label=f'AUC = {auc:.2f}')
plt.plot([0, 1], [0, 1], 'k--')  # Diagonal line for random classifier
plt.xlabel('False Positive Rate')
plt.ylabel('True Positive Rate')
plt.title('ROC Curve')
plt.legend(loc='lower right')
plt.savefig('./figure.pdf')
```
